# Routine use of oral iron for people with heart failure and iron deficiency in primary care; retrospective cohort study

**DOI:** 10.64898/2026.06.14.26355620

**Authors:** Vijay Maharajan, Nicholas R Jones, Clare Bankhead, Innocent Erone, Sarah Haynes, Alvin Katumba, Christien Ka Hou Li, Suzanne Maynard, Noemi BA Roy, Akshay Shah, Simon J Stanworth, Margaret Smith, Cynthia Wright Drakesmith

**Affiliations:** Cardio-Oncology Service, Royal Brompton Hospital Centre of Excellence, Guy’s and St Thomas’ NHS Foundation Trust, London, UK; University of Oxford Nuffield Department of Primary Care Health Sciences, Oxford, UK; Department of Cardiology, Oxford University Hospital, Oxford, UK; University of Oxford, Nuffield Department of Women’s and Reproductive Health, Oxford, UK; University of Oxford, Radcliffe Department of Medicine, Oxford, UK; Department of Anaesthesia, Hammersmith Hospital, Imperial College Healthcare NHS Trust, London, UK; Nuffield Department of Clinical Neurosciences, University of Oxford, Oxford, UK; Department of Haematology, Oxford University Hospitals NHS Trust, Oxford, UK; NHSBT, Oxford, UK; Department of Transfusion Medicine, NHS Blood and Transplant, Oxford, UK

## Abstract

**Aims:** Iron deficiency is common among people with heart failure and associated with morbidity and mortality. While intravenous iron improves clinical outcomes, oral iron continues to be prescribed in routine practice despite limited evidence of benefit.

**Methods:** We completed a retrospective primary care cohort study (2016 to 2021) to investigate the proportion of people with an incident diagnosis of heart failure who had iron deficiency identified (defined as ferritin <100µg/L) and subsequently received a first prescription for oral iron within 12 months. Multivariable logistic regression was used to report the odds ratio (OR) of receiving oral iron in relation to key demographic covariates and co-morbidities.

**Results:** Among 105,749 people with an incident diagnosis of heart failure, 35,688 underwent a ferritin test within the first year of whom 11,237 had iron deficiency and no prior prescription for oral iron. Of these, 2,734 (24.3%) were subsequently prescribed oral iron. Increasing age (OR per 10-year increase 1.14, 95%CI: 1.10-1.19), Asian ethnicity (1.33, 1.08-1.64), cirrhosis (2.01, 1.29-3.14) and diabetes (1.36, 1.24-1.49) were associated with increased odds of receiving oral iron. Among 1,357 (49.6%) people who had their ferritin level re-tested, the median change was 26 µg/L (interquartile range 7 to 61) among people who were prescribed oral iron compared to 4 µg/L (IQR −9 to 34) among people not prescribed oral iron.

**Conclusions:** One in four individuals with heart failure and low ferritin received oral iron replacement, despite this not being recommended in international guidelines. Treatment could be improved and standardised in primary care.

## Introduction

Iron deficiency is highly prevalent among people with heart failure, affecting up to 50% of patients in some studies.^1^ It is independently associated with a greater symptom burden, including fatigue and dyspnoea, and an overall reduction in quality of life, even in the absence of anaemia.^2^ Beyond symptomatic impact, iron deficiency in heart failure is also linked with a poorer prognosis and with markers of disease severity, including elevated N-terminal pro-B-type natriuretic peptide (NT-proBNP) levels and lower left ventricular ejection fraction (LVEF).^3^ ^4^

International guidelines recommend that people with heart failure should be checked for iron deficiency to inform both prognosis and treatment with intravenous iron.^5–7^ For example, since 2016 the European Society of Cardiology (ESC) guidelines have recommended testing ferritin and transferrin saturations (TSAT) in patients newly diagnosed with heart failure.^8^ Randomised control trials have demonstrated that intravenous iron replacement for those with heart failure and reduced ejection fraction (HFrEF) and iron deficiency can increase serum iron levels, improve quality of life and reduce hospitalisations for heart failure.^9–11^ These studies did not demonstrate a reduction in all-cause mortality, although they may have been under-powered to detect this.^12^ ^13^ There was a class IIA recommendation for intravenous iron replacement in people with iron deficiency and heart failure in the ESC 2016 guidelines, upgraded to a class IA recommendation in 2023 on the basis of the evolving evidence base.^7^ ^8^

Guidelines explicitly recommend intravenous iron replacement because the impact of oral iron in people with heart failure is unclear. Although modest improvements in ferritin have been reported, there is uncertainty regarding effects on improving symptom scores, even among selected patient sub-groups.^14^ ^15^ However, clinicians may continue to prescribe oral iron for people with heart failure given it is cheap and readily available, despite challenges with tolerance, efficacy and adherence. The extent to which this is happening is unclear, meaning there is uncertainty regarding current practice and implementation, including which patient groups are most likely to be prescribed oral iron. Previous studies of oral iron for heart failure were done in relatively small and selected populations with short-term follow-up so cannot report on national prescribing patterns. They may also under-estimate the potential benefit of oral iron across broader populations. If oral iron were effective it would provide a lower cost and accessible alternative to intravenous treatment.

In this study we aim to report on the proportion of people with an incident diagnosis of heart failure and a ferritin < 100 µg/L who received oral iron replacement in routine primary care, the patient factors associated with receiving treatment and the subsequent change in serum ferritin over time. The results can help inform to what extent current practice is compliant with the recommendation for intravenous rather than oral iron replacement and explore any potential treatment response to oral iron in a large primary care cohort.

## Methods

### Study design and setting

This was a retrospective cohort study conducted using United Kingdom (UK) primary care data from the Clinical Practice Research Datalink (CPRD) Aurum between January 2016 and March 2021. The Aurum database includes over 40 million patient records from 1,800 GP practices.^16^ Access to the data is provided with permission from the CPRD’s Expert Review Committee (eRAP number 22_001873) and under the umbrella approval of the NHS Research Ethics Committee for use of anonymised patient data. The manuscript is reported in accordance with The REporting of studies Conducted using Observational Routinely-collected health Data (RECORD) Statement (Supplementary Table 1).^17^

### Participants

We included patients if they were aged ≥18 years and had an incident diagnostic code for heart failure in their primary care record (EHR). This analysis was restricted to individuals who met the ESC criteria for iron deficiency within the first year following their heart failure diagnosis. Iron deficiency was defined as a serum ferritin <100 µg/L measured within 90 days prior to the incident heart failure diagnosis or up to 12 months after, even in the absence of anaemia.^6^ ^18^ The short window prior to a diagnosis was included as tests may be done as part of the heart failure diagnostic work-up without then being repeated once the diagnosis was confirmed. We excluded patients with less than 12 months of follow-up data as this would include individuals who were close to end of life where iron testing and treatment may not be in the patient’s best interests. We also excluded patients with implausible ferritin values, such as a negative result or ferritin greater than 100,000 µg/L.

### Variables

The primary outcome was a first primary care prescription for any type of oral iron within the first year following an incident heart failure diagnosis among individuals who had never previously been prescribed oral iron in primary care prior to the heart failure diagnosis. The secondary outcomes were the factors associated with a prescription for oral iron and the subsequent change in serum ferritin levels among those treated. A code list was created to identify oral iron prescriptions in the primary care record, including ferrous sulphate, ferrous fumarate and ferrous gluconate at any dose. These prescriptions were extracted along with the date the prescription was dispensed, with the date of the first prescription following heart failure diagnosis defined as the start date of oral iron therapy.

Ferritin interpretation can be complicated by the presence of other conditions with an inflammatory component that typically raise serum ferritin, but may also be associated with iron deficiency. To investigate this, through expert Delphi consensus a multidisciplinary team of clinicians including both GPs and haematologists prioritised conditions based on prevalence and relevance to iron testing and interpretation of serum ferritin.^19^ This resulted in a list of 19 chronic inflammatory or systemic conditions included in our analysis: chronic kidney disease (CKD, stages 3–5), chronic obstructive pulmonary disease (COPD), dementia, osteoarthritis, Parkinson’s disease, rheumatoid arthritis, stroke, bronchiectasis, multiple sclerosis (MS), polycystic ovary syndrome (PCOS), endometriosis, liver cirrhosis, gastro-oesophageal reflux disease (GORD), irritable bowel syndrome (IBS), allergic rhinitis, ulcerative colitis, Crohn’s disease, coeliac disease, and diabetes mellitus (both combined and separated into type 1 and type 2 diabetes). Patients were categorised as having a condition if a diagnostic code was present in their primary care record prior to the incident heart failure date. We also collected demographic data on age, sex, ethnicity (reported into five categories based on the Census 2021 approach.^20^) and geographic region, with the latter based upon CPRD’s practice-level geographic variable.

### Statistical analysis

The cohort is summarised using descriptive statistics, stratified by whether patients with iron deficiency received an incident prescription for oral iron within one year of heart failure diagnosis. Counts and percentages are reported for categorical variables and means with standard deviation (SD) or median with interquartile ranges (IQR) for continuous data, as appropriate. We report the odds ratio (OR) of receiving a prescription for oral iron in relation to relevant patient characteristics using a multivariable logistic regression that was adjusted for age, sex, ethnicity and National Health Service (NHS) region. Each MLTC was subsequently tested individually for association with treatment response. We undertook a sensitivity analysis including people with incident heart failure who had previously been prescribed oral iron before the heart failure diagnosis date.

Among the individuals who had two ferritin results, we report the median change in ferritin between the first and second test, comparing between people who were or were not prescribed oral iron. Linear regression was used to assess subsequent changes in ferritin between the first and second measurements adjusted for age, sex and ethnicity. Change in ferritin was log-transformed given it was not normally distributed. We also reported recovery from iron deficiency as a binary outcome, using logistic regression to report the odds of ferritin recovering to ≥100 µg/L at follow-up, representing the ESC treatment threshold. We report the mean and SD for the time between tests overall and comparing between people whose ferritin level improved to ≥100 µg/L on the second test to those in whom it didn’t, to explore whether time to repeat testing was important in explaining ferritin recovery to above the ESC threshold for defining iron deficiency. A Mann Whitney U test was used to assess for a significant difference in changes in ferritin between groups. To investigate whether differences in improvement in ferritin tests in relation to time were different in people who were or were not prescribed oral iron we also performed linear regression of the log-transformed change in ferritin value between tests that was adjusted for age, sex and ethnicity and which included an interaction term for an incident prescription for oral iron and the time in weeks between two ferritin measurements. The exponential of this result is reported including the 95% confidence interval.

Age, sex and practice registration had to be complete for patients to be included in the cohort. Diagnostic and test data were assumed to be complete, meaning missing data were confined to ethnicity and geographic region. Both were considered to be missing not at random (MNAR), making multiple imputation inappropriate.^21^ We instead created separate ‘missing’ indicators for each of these variables, which were included in the regression models. All statistical analyses were conducted in Stata 18, following data extraction in SQL and pre-processing in Python.

## Results

Within the database we identified 105,749 people with an incident diagnosis of heart failure between January 2016 and March 2021. Of these, 35,688 (33.7%) individuals had a ferritin test within the first year of their diagnosis with heart failure (Figure 1). There were 11,237 (31.5% of those tested) people who met the ESC criteria for iron deficiency based on this test, and who had no prior prescription for iron in their primary care record. Of these, 2,734 (24.3%) received a primary care prescription for oral iron.

**Figure 1:**
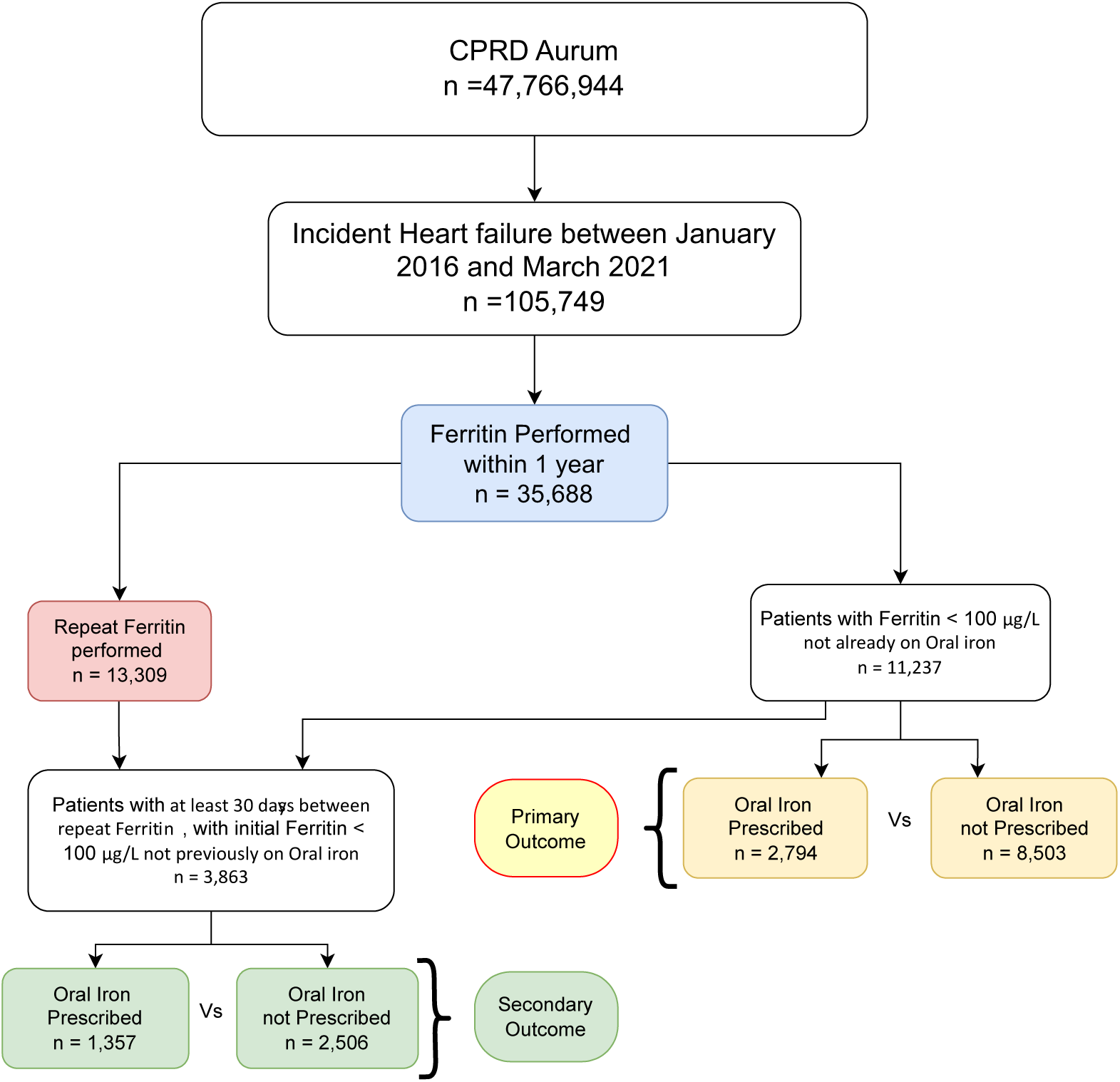
Flow diagram of study cohort From 47.8 million patients in CPRD Aurum, 105,749 with incident heart failure were identified. Subsequent exclusions and subgrouping are shown for analyses of ferritin testing, repeat testing, treatment, and effectiveness.

The majority of people who had incident heart failure with iron deficiency in the cohort were older (e.g. 49.6% (n=5,570) aged 75 to 89 years), of white ethnicity (n=8,961, 79.7%) and had multiple other long-term health conditions, of which the most prevalent were osteoarthritis (n=5,028, 44.7%) and diabetes (n=4,066, 36.2%) (Table 1). There were similar numbers of males (5,321, 47.4%) and females (5,916, 52.6%) with incident heart failure and iron deficiency.

**Table 1.**
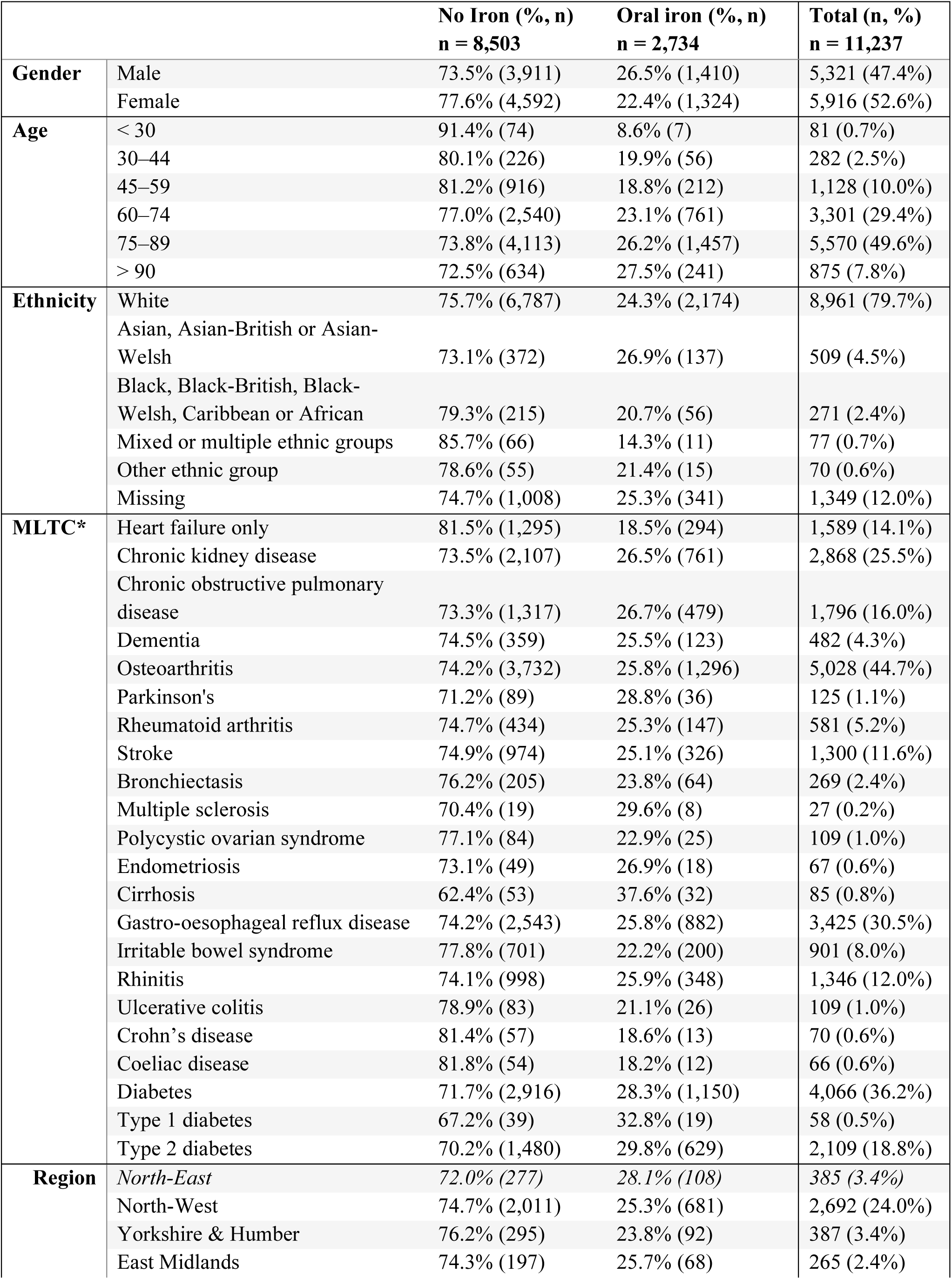
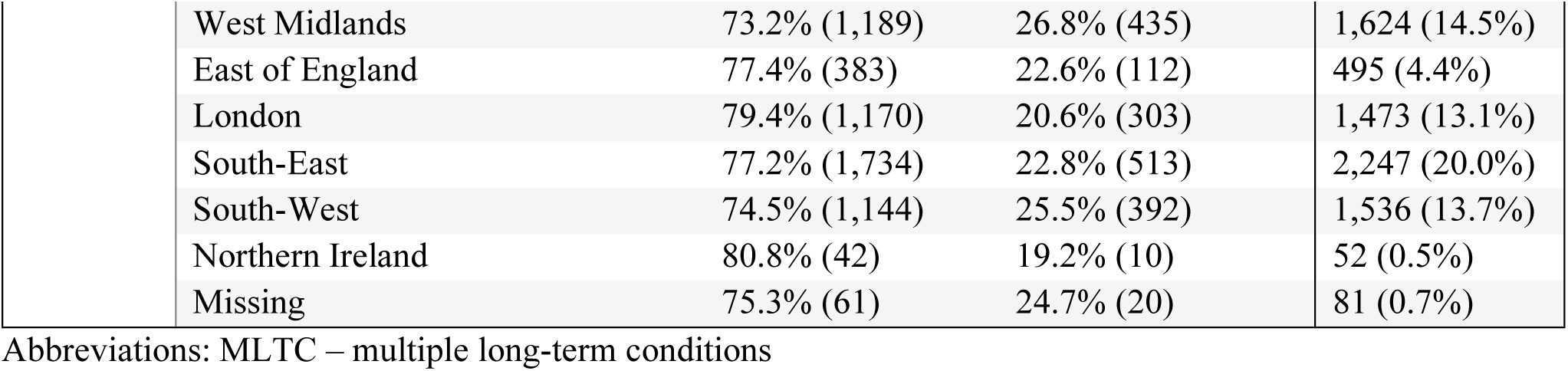
Baseline demographic and clinical characteristics of patients with heart failure and iron deficiency, stratified by whether oral iron was prescribed

|  |  | **No Iron (% , n)**<br>**n = 8,503** | **Oral iron (% , n)**<br>**n = 2,734** | **Total (n, %)**<br>**n = 11,237** |
| --- | --- | --- | --- | --- |
| **Gender** | Male | 73.5% (3,911) | 26.5% (1,410) | 5,321 (47.4%) |
|  | Female | 77.6% (4,592) | 22.4% (1,324) | 5,916 (52.6%) |
| **Age** | < 30 | 91.4% (74) | 8.6% (7) | 81 (0.7%) |
|  | 30–44 | 80.1% (226) | 19.9% (56) | 282 (2.5%) |
|  | 45–59 | 81.2% (916) | 18.8% (212) | 1,128 (10.0%) |
|  | 60–74 | 77.0% (2,540) | 23.1% (761) | 3,301 (29.4%) |
|  | 75–89 | 73.8% (4,113) | 26.2% (1,457) | 5,570 (49.6%) |
|  | > 90 | 72.5% (634) | 27.5% (241) | 875 (7.8%) |
| **Ethnicity** | White | 75.7% (6,787) | 24.3% (2,174) | 8,961 (79.7%) |
|  | Asian, Asian-British or Asian-Welsh | 73.1% (372) | 26.9% (137) | 509 (4.5%) |
|  | Black, Black-British, Black-Welsh, Caribbean or African | 79.3% (215) | 20.7% (56) | 271 (2.4%) |
|  | Mixed or multiple ethnic groups | 85.7% (66) | 14.3% (11) | 77 (0.7%) |
|  | Other ethnic group | 78.6% (55) | 21.4% (15) | 70 (0.6%) |
|  | Missing | 74.7% (1,008) | 25.3% (341) | 1,349 (12.0%) |
| **MLTC\*** | Heart failure only | 81.5% (1,295) | 18.5% (294) | 1,589 (14.1%) |
|  | Chronic kidney disease | 73.5% (2,107) | 26.5% (761) | 2,868 (25.5%) |
|  | Chronic obstructive pulmonary disease | 73.3% (1,317) | 26.7% (479) | 1,796 (16.0%) |
|  | Dementia | 74.5% (359) | 25.5% (123) | 482 (4.3%) |
|  | Osteoarthritis | 74.2% (3,732) | 25.8% (1,296) | 5,028 (44.7%) |
|  | Parkinson's | 71.2% (89) | 28.8% (36) | 125 (1.1%) |
|  | Rheumatoid arthritis | 74.7% (434) | 25.3% (147) | 581 (5.2%) |
|  | Stroke | 74.9% (974) | 25.1% (326) | 1,300 (11.6%) |
|  | Bronchiectasis | 76.2% (205) | 23.8% (64) | 269 (2.4%) |
|  | Multiple sclerosis | 70.4% (19) | 29.6% (8) | 27 (0.2%) |
|  | Polycystic ovarian syndrome | 77.1% (84) | 22.9% (25) | 109 (1.0%) |
|  | Endometriosis | 73.1% (49) | 26.9% (18) | 67 (0.6%) |
|  | Cirrhosis | 62.4% (53) | 37.6% (32) | 85 (0.8%) |
|  | Gastro-oesophageal reflux disease | 74.2% (2,543) | 25.8% (882) | 3,425 (30.5%) |
|  | Irritable bowel syndrome | 77.8% (701) | 22.2% (200) | 901 (8.0%) |
|  | Rhinitis | 74.1% (998) | 25.9% (348) | 1,346 (12.0%) |
|  | Ulcerative colitis | 78.9% (83) | 21.1% (26) | 109 (1.0%) |
|  | Crohn's disease | 81.4% (57) | 18.6% (13) | 70 (0.6%) |
|  | Coeliac disease | 81.8% (54) | 18.2% (12) | 66 (0.6%) |
|  | Diabetes | 71.7% (2,916) | 28.3% (1,150) | 4,066 (36.2%) |
|  | Type 1 diabetes | 67.2% (39) | 32.8% (19) | 58 (0.5%) |
|  | Type 2 diabetes | 70.2% (1,480) | 29.8% (629) | 2,109 (18.8%) |
| **Region** | *North-East* | *72.0% (277)* | *28.1% (108)* | *385 (3.4%)* |
|  | North-West | 74.7% (2,011) | 25.3% (681) | 2,692 (24.0%) |
|  | Yorkshire & Humber | 76.2% (295) | 23.8% (92) | 387 (3.4%) |
|  | East Midlands | 74.3% (197) | 25.7% (68) | 265 (2.4%) |

|  |  |  |  |
| --- | --- | --- | --- |
| West Midlands | 73.2% (1,189) | 26.8% (435) | 1,624 (14.5%) |
| East of England | 77.4% (383) | 22.6% (112) | 495 (4.4%) |
| London | 79.4% (1,170) | 20.6% (303) | 1,473 (13.1%) |
| South-East | 77.2% (1,734) | 22.8% (513) | 2,247 (20.0%) |
| South-West | 74.5% (1,144) | 25.5% (392) | 1,536 (13.7%) |
| Northern Ireland | 80.8% (42) | 19.2% (10) | 52 (0.5%) |
| Missing | 75.3% (61) | 24.7% (20) | 81 (0.7%) |

Abbreviations: MLTC – multiple long-term conditions

### Predictors of incident oral iron prescribing

The proportion of people with an incident prescription for oral iron following their heart failure diagnosis increased with age (adjusted OR per 10 year increase 1.14, 95%CI 1.10 to 1.19) (Table 2), with only 8.6% (n=7) of those aged under 30 years receiving a prescription compared to 26.2% (n=1,457) of those aged 75 to 89 years and 27.5% (n=241) of those aged 90 years or older (Table 1). A slightly greater proportion of males (n=1,410, 26.5%) than females (n=1,324, 22.4%) were first prescribed iron (OR 1.28, 95%CI: 1.18 to 1.40) following a diagnosis of heart failure (Table 2). Prescribing was largely similar between different ethnic groups though lowest among those of mixed or multiple ethnic groups (14.3%, n=11) and highest among the Asian, Asian-British or Asian-Welsh population (26.9%, n=137) who also had the highest odds ratio for a prescription (OR 1.33, 95%CI: 1.08 to 1.64 compared to white ethnicity). Comparing within geographic regions, the highest proportion of people prescribed oral iron were those in the North-East of England (28.1%, n=108) (OR 1.52, 95%CI: 1.17 to 1.97), followed by the West Midlands (26.8%, n=435) (OR 1.38, 95%CI: 1.16 to 1.64) and the lowest proportion those in Northern Ireland (19.2%, n=10), followed by London (20.6%, n=303) (OR – reference group).

**Table 2.** Predictors of oral iron prescribing in patients with heart failure and confirmed iron deficiency, stratified by long-term condition.

|  |  | **Odds Ratio** | **95% CI** | **P-value** |
| --- | --- | --- | --- | --- |
| **Age (per 10 years)** |  | 1.14 | (1.10 - 1.19) | <0.001 |
| **Gender ref: female** | Male | 1.28 | (1.18 - 1.40) | <0.001 |
| **Ethnicity ref: White** | Asian, Asian British or Asian Welsh | 1.33 | (1.08 - 1.64) | 0.008 |
|  | Black, Black British, Black Welsh, Caribbean or African | 1.00 | (0.73 - 1.36) | 0.993 |
|  | Mixed or multiple ethnic groups | 0.64 | (0.33 - 1.21) | 0.171 |
|  | Other ethnic group | 1.05 | (0.59 - 1.87) | 0.869 |
|  | Missing | 1.02 | (0.89 - 1.17) | 0.741 |
| **Region ref: London** | North-East | 1.52 | (1.17 - 1.97) | 0.002 |
|  | North-West | 1.31 | (1.11 - 1.54) | 0.001 |
|  | Yorkshire and The Humber | 1.17 | (0.89 - 1.53) | 0.259 |
|  | East Midlands | 1.3 | (0.96 - 1.77) | 0.103 |
|  | West Midlands | 1.38 | (1.16 - 1.64) | <0.001 |
|  | East of England | 1.09 | (0.84 - 1.40) | 0.522 |
|  | South-East | 1.13 | (0.95 - 1.34) | 0.164 |
|  | South-West | 1.31 | (1.00 - 1.70) | 0.037 |
|  | Northern Ireland | 1.02 | (0.51 - 2.07) | 0.944 |
|  | Missing | 1.23 | (0.73 - 2.07) | 0.445 |
| **Condition** | Heart failure only | 0.75 | (0.66 - 0.87) | <0.001 |
|  | Chronic kidney disease | 1.06 | (0.96 - 1.18) | 0.24 |
|  | Chronic obstructive pulmonary disease | 1.13 | (1.00 - 1.26) | 0.045 |
|  | Dementia | 0.97 | (0.78 - 1.20) | 0.763 |
|  | Osteoarthritis | 1.07 | (0.98 - 1.18) | 0.126 |
|  | Parkinson's | 1.16 | (0.78 - 1.71) | 0.466 |
|  | Rheumatoid arthritis | 1.07 | (0.88 - 1.30) | 0.497 |
|  | Stroke | 0.91 | (0.84 - 1.10) | 0.56 |
|  | Bronchiectasis | 0.92 | (0.69 - 1.23) | 0.581 |
|  | Multiple sclerosis | 1.5 | (0.65 - 3.44) | 0.343 |
|  | Polycystic ovarian syndrome | 1.25 | (0.79 - 1.96) | 0.346 |
|  | Endometriosis | 1.47 | (0.85 - 2.55) | 0.166 |
|  | Cirrhosis | 2.01 | (1.29 - 3.14) | 0.002 |
|  | Gastro-oesophageal reflux disease | 1.1 | (1.00 - 1.21) | 0.051 |
|  | Irritable bowel syndrome | 0.95 | (0.81 - 1.12) | 0.548 |
|  | Rhinitis | 1.08 | (0.95 - 1.23) | 0.255 |
|  | Ulcerative colitis | 0.82 | (0.52 - 1.30) | 0.401 |
|  | Crohn's disease | 0.75 | (0.41 - 1.37) | 0.348 |
|  | Coeliac disease | 0.68 | (0.36 - 1.27) | 0.227 |
|  | Diabetes | 1.36 | (1.24 - 1.49) | <0.001 |
|  | Type 1 diabetes | 1.78 | (1.02 - 3.10) | 0.042 |
|  | Type 2 diabetes | 1.37 | (1.24 - 1.53) | <0.001 |

*Multivariable logistic regression adjusted for age, sex, ethnicity and NHS region.*

A higher proportion of people with certain co-morbidities received an oral iron prescription including people with chronic kidney disease (26.5%, n=761), chronic obstructive pulmonary disease (26.7%, n=479), Parkinson’s disease (28.8%, n=36), cirrhosis (37.6%, n=32) and type 1 diabetes (32.8%, n=19). However, the odds ratio was only statistically significant for cirrhosis (OR 2.01, 95%CI: 1.29 to 3.14) and diabetes (OR 1.36, 95%CI: 1.24 to 1.49) when compared to people with heart failure but no other long-term conditions. Only 18.5% (n=294) of patients with heart failure alone and iron deficiency received oral iron.

In a sensitivity analysis including 8,364 patients who had a prevalent prescription for oral iron before their heart failure diagnosis, females had higher odds of a prescription for oral iron in the year following their heart failure diagnosis than males (OR 0.83, 95%CI 0.78 to 0.88) (Supplementary Table 2) reflecting a higher baseline prevalence of oral iron use among women compared with men prior to heart failure diagnosis, among patients who met ESC criteria for iron deficiency (46.6% [5,163/11,079] vs 37.6% [3,201/8,522]). Other results were in line with the main analysis, although the odds ratio for oral iron increased significantly among both Asian (OR 2.42, 95% CI: 2.12–2.75) and Black patients (OR 1.51, 95% CI: 1.26–1.82 (Supplementary Table 2))

### Treatment response to oral iron

Of the 35,688 people with incident heart failure and an initial ferritin test, 13,309 had a second ferritin test within the next year (Figure 1). Amongst these, 3,863 (29.0%) met ESC-defined criteria for iron deficiency and had no prior oral iron exposure (Table 3). There were 1,357 (35.1%) individuals who received a first prescription for oral iron between their first and second ferritin tests (Supplementary Table 3). The median change in ferritin between the first and second test was greater in the group who received oral iron (26 µg/L, IQR 7 to 61) compared to the the group who did not receive oral iron (4 µg/L, IQR −9 to 34) (Table 3). Linear regression confirmed that oral iron use was associated with increased ferritin levels after adjustment for age, sex, and ethnicity (OR = 1.22, 95% CI: 1.09 to 1.36). However, the proportion of people whose second ferritin test improved to ≥100 µg/L was similar between who did or did not receive a prescription for oral iron (Table 3). Similarly, in a logistic regression analysis, oral iron did not improve the odds of a second ferritin test ≥100 µg/L (OR 0.92, 95% CI: 0.78 to 1.08). This ferritin value (≥100 µg/L) reflects the ESC threshold for iron deficiency based on ferritin alone, demonstrating that the majority of people continued to have iron deficiency by this measure, whether or not they received oral iron.^7^

**Table 3.** Changes in ferritin values in patients with heart failure and confirmed iron deficiency who received at least two ferritin tests during the study period, comparing between people who did or did not receive a prescription for oral iron

|  | People who received a prescription for oral iron | People who did not receive a prescription for oral iron |
| --- | --- | --- |
| Number of participants who received two or more ferritin tests within study period | 1357 | 2506 |
| Change in ferritin between first and second test (median, interquartile range) | 26µg/L, 7 to 61µg/L | 4µg/L, -9 to 34µg/L |
| Linear regression for increased ferritin level between tests (odds ratio, 95% confidence interval) | 1.22 (1.09 to 1.36) | Reference |
| People whose ferritin improved to ≥100 µg/L by second test (number, percentage) | 299 (22.0%) | 584 (23.3%) |
| Linear regression for ferritin improving to ≥100 µg/L by second test (odds ratio, 95% confidence interval) | 0.92 (0.78 to 1.08) | Reference |

Among the people who had multiple ferritin tests, the mean time between ferritin test dates was 30.6 weeks (SD 14.0). In regression analysis adjusted for age, sex and ethnicity, a longer interval between the two measurements of serum ferritin was associated with greater increases in ferritin levels (OR = 1.02 per additional week between tests, 95%CI 1.01 to 1.02). The mean time between ferritin tests among people whose ferritin recovered to ≥100 µg/L was 32.6 weeks (SD 13.7), compared to a mean time between tests of 30.0 weeks (SD 14.0) among people whose ferritin remained <100 µg/L. In an adjusted regression, the interaction between oral iron and the time between ferritin tests was not significant (OR 0.76, 95%CI: 0.53 to 1.07), suggesting that a longer time period between ferritin tests was similarly associated with an increased likelihood of the ferritin recovering to ≥100 µg/L, irrespective of whether a patient was prescribed oral iron.

## Discussion

We report a large-scale retrospective cohort study of oral iron prescribing among people with heart failure in primary care. Only a third of people with incident heart failure underwent ferritin testing in the first year and a third of them had a serum ferritin <100 µg/L, meeting ESC criteria for iron deficiency. Of these people we found that one in four received a first prescription for oral iron within the subsequent twelve months, despite this not being recommended treatment.^5^ ^6^ The likelihood of receiving oral iron was higher with increasing age, among people of Asian ethnicity and among people with certain long-term comorbidities, particularly cirrhosis or diabetes. Although oral iron was associated with a statistically significant increase in serum ferritin among people who had this re-tested, it is unclear whether this is clinically meaningful and there was no significant association between oral iron prescription and people becoming replete of iron, defined as a serum ferritin ≥100 µg/L over a mean period of 30 weeks. Our results highlight important gaps between international guideline recommendations and current clinical practice given the low proportion of people with incident heart failure who are being tested for heart failure and the relatively high proportion who are found to have iron deficiency and subsequently treated with oral iron.

Our findings with respect to response to oral iron as treatment for iron deficiency in people with heart failure are in line with previous randomised trials that have demonstrated only modest improvements in serum ferritin through this treatment approach.^14^ ^22^ For example, in the IRONOUT trial, oral iron was associated with a 11.3ng/mL (95%CI: −0.3 to 22.9ng/mL) increase in serum ferritin and a 3.3% (95%CI: 1.1 to 5.4%) increase in TSAT levels, but no improvement in the primary outcome of peak oxygen consumption at 16 weeks.^14^ Similarly, in a recent systematic review oral iron treatment for people with heart failure and iron deficiency did not improve exercise capacity measured by a 6 minute walk test and only resulted in small, non-statistically significant improvements in serum ferritin or TSAT.^23^ Within our cohort, oral iron was also associated with a statistically significant increase in serum ferritin (+24 µg/L (p < 0.001), but this was small and we did not find a significant difference in the proportion of individuals who were iron replete, defined by a ferritin above the ESC threshold of 100 µg/L.

Within IRONOUT it was noted that there was an association between higher baseline hepcidin levels and a decreased probability of becoming iron replete through oral iron replacement.^14^ In states of chronic inflammation, as is common in heart failure, hepcidin is upregulated, leading to functional iron deficiency despite preserved stores.^24^ The limited response to oral iron may therefore partly be explained by hepcidin inhibiting its absorption.^25^ Other potential factors include gut oedema limiting iron absorption, poor adherence with oral iron or increased blood loss among patients treated with antiplatelets or anticoagulants.^1^ ^26^

### Strengths and limitations

Our study uses a large primary care dataset that is representative of the general population in England. The study was primarily designed to report on the proportion of people with heart failure who were prescribed oral iron and these data along with ferritin tests are generally well coded.

However, there is significant uncertainty when interpreting the response to oral iron because of the lack of data on the type of oral iron used, dosing schedules, adherence to treatment and treatment duration. Extracting dosage information from CPRD is technically challenging and was beyond the scope of this analysis. It is possible that patients prescribed higher doses of oral iron might have had a more substantial improvement in ferritin. However, very high doses of oral iron can reduce absorption and previous trials have demonstrated similar improvement in ferritin levels with alternate day compared to daily iron replacement among people with iron deficiency.^27^ ^28^ In the IRONOUT study the dose of oral iron was approximately 15 times higher than that used in some previous studies of intravenous iron, suggesting the lack of response to oral iron is not a consequence of inadequate dosing.^14^ ^29^ Some patients may have had tests or treatment for iron deficiency done in secondary care and this would have been missed from our analysis.

We also did not collect data on patients referred for intravenous iron replacement and this could have resulted in some patients who did not receive oral iron having a significant improvement in iron stores. Such a change would dilute any apparent treatment effect of oral iron and it was notable that the ferritin levels on average improved even in the group who were not prescribed oral iron.

Ferritin values may also have been affected by other clinical factors not captured in our analysis, such as treatment for inflammatory conditions. We cannot tell the indication for ferritin testing from the healthcare record and there may also have been selection bias in terms of which patients were selected for both initial and repeat ferritin testing that could have impacted the results. This makes the comparative change in ferritin between the groups who did and did not receive oral iron difficult to interpret. Nevertheless, our findings demonstrate that around a quarter of people with incident heart failure and iron deficiency were prescribed oral iron, despite this not being recommended in international guidelines and that only a low proportion of those given oral iron became iron replete during follow-up.

Ferritin is a non-specific test for iron deficiency and the level can be altered in other situations, such as patients with acute infection or inflammatory conditions. National Institute for Health and Care Excellence (NICE) guidelines in the UK recommend that TSAT is also measured when considering iron deficiency as well as haemoglobin to check for anaemia.^5^ However, a feasibility check within Aurum demonstrated less than 5% of patients within the cohort had a TSAT test. We therefore limited our analysis to ferritin given this is the most frequently used test and forms the basis for the ESC definition of iron deficiency but recognise that this approach does risk misclassifying some individuals.^7^ Ideally other indicators of response to oral iron would have been included in our analysis, including haemoglobin and functional tests that measure exercise capacity or symptom scores.

Heart failure is a heterogeneous condition, but one approach to classification is by LVEF, which typically have different underlying aetiologies, which might pre-dispose to different response to iron replacement. The data supporting the use of intravenous iron has predominantly been generated among people with HFrEF and so categories of heart failure may have impacted on testing and treatment in practice. However, LVEF is poorly coded in CPRD meaning it was not possible to report results on this basis.

### Implications for research and practice

Only a third of people with an incident diagnosis of heart failure undergo ferritin testing within the subsequent year, yet around a third of patients tested were found to be iron deficient with a serum ferritin <100 µg/L. These results demonstrate the high prevalence of iron deficiency in the heart failure population and mean that there is an opportunity to substantially increase testing and identification of iron deficiency in primary care. However, among those tested a quarter of patients were prescribed oral iron replacement despite this not being recommended in international heart failure guidelines. Potential reasons for this include limited access to intravenous iron treatment, a lack of knowledge of the guidelines or patient preference to trial oral treatments first. Public health and educational initiatives might target people with heart failure and primary care clinicians respectively with the aim of improving awareness of the recommendations for intravenous iron treatment to try to address this treatment gap. There is a lack clear guidance around when repeat ferritin or TSAT testing should be performed and this could help provide clarity to clinicians.

Although we did not find evidence of significant clinical benefit through oral iron replacement, further prospective studies could explore the potential benefit of alternate day dosing or novel oral iron formulations, which may both lead to better absorption through less significant or sustained increases in hepcidin levels.^30^ ^31^

## Conclusions

Our analysis demonstrates a significant gap between current international guidelines and clinical practice relating to the identification and management of people with iron deficiency and heart failure. A quarter of the cohort with heart failure who met the criteria for iron deficiency were prescribed oral iron treatment, despite this not being recommended in international guidelines.

Increasing age, Asian ethnicity and co-morbidity with cirrhosis or diabetes were associated with receiving oral iron. Prescription of oral iron was associated with a small increase in serum ferritin but it is not clear if this is clinically significant and, on average, it did not lead to resolution of iron deficiency, supporting the current approach of referring patients for intravenous iron replacement in this situation. Further work is needed to raise awareness of the guidance around iron deficiency in people with heart failure and also improvements in access to services providing intravenous iron replacement at scale in the community.

## Data Availability

The data used in this study were obtained from the Clinical Practice Research Datalink (CPRD) under licence and cannot be shared directly by the authors. Access to CPRD data is subject to approval through the CPRD Research Data Governance process. Further information on data access is available from CPRD.

https://www.cprd.com/access-data

## Acknowledgements

This study is based on data from the Clinical Practice Research Datalink (CPRD) obtained under licence from the UK Medicines and Healthcare products Regulatory Agency. The data is provided by patients and collected by the NHS as part of their care and support. The interpretation and conclusions contained in this study are those of the author alone. This work was carried out under approved study protocol 22_001873. This project is supported (in part) by the UK NIHR Blood and Transplant Research Unit in Data Driven Transfusion Practice (NIHR203334).

## Data availability

The data used for this analysis was provided under license by the Clinical Practice Research Datalink (CPRD) for the purpose of this study alone and the authors are not able to share the data. However, requests to access the same data can be made directly to CPRD via https://www.cprd.com/access-data.

